# An epidemiological study of migraine headaches among the staff of Shahid Beheshti Hospital in Kashan, Iran, and related factors

**DOI:** 10.1101/2022.07.21.22277660

**Authors:** Seyed-amir abbas ahadiat, Zeinab Hosseinian, Ebrahim kouchaki

## Abstract

**Introduction:** Migraine, as one of the most common types of disability, has a high prevalence with significant effects on a person’s living conditions. This study aimed to estimate the prevalence and evaluate the clinical characteristics of migraine patients in Shahid Beheshti Hospital in Kashan.

**Method:** This study was performed cross-sectionally. All hospital staff (1125 people) were studied in 2019. Based on a designed questionnaire, the prevalence of migraine headaches and their relationship with occupations, marital status, age, gender, work shift, hours of sleep per day, as well as accompanying symptoms, were analyzed using analytic software.

**Results:** The prevalence of migraine among hospital staff was 12.3% (n=136), and nurses with 14.5% had the highest majority, and faculty members with 9.7% had the lowest prevalence (15.5% in females and 8.5% in males). 89.7% of people had unilateral headache. The maximum number of migraine attacks per month was five (32.23%), and the most common associated symptoms were anorexia(p-value=0.048), nausea(p-value=0.003), and vomiting(p-value=0.97), respectively. There was a significant relationship between migraine with sleep hours per day(p-value=0.012), age(p-value=0.035), and marital status (p-vaue=0.001).

**Conclusion:** findings relating to severe headaches and subsequent loss of work are important in realizing the social impact of this condition and the need for prevention and treatment strategies. In other words, primary prevention, diagnosis, and treatment should receive more attention.

## introduction

Migraine is one of the commonest disabling chronic neurological diseases in the world (1). Females are more likely to suffer from migraines than men. Migraine attacks are most common between the ages of 30 and 39 in western countries and Turkey (2); while there is no exact prevalence of migraines worldwide, that can vary by race and geography. (3). According to a WHO report, the incidence of migraine in America and Europe in adults was estimated at about 10–15% per year. in the pandemic covid-19, migraine patients will have a difficult time finding optimum care due to access problems, social isolation, and increased stress, also the annual cost of treatment and individuals’ being absent due to migraine headaches is about 50 billion dollars(4-5)

According to a study in 2007 in Iran, 8.1% of the students were suffering from migraine headaches, and it was more common among single students(6). According to a systematic review and meta-Analysis by Farhadi et al, migraine prevalence in Iran was estimated at 14%, similar to or higher than worldwide. (7).A similar study among hospital staff in shiraz determined prevalence of migraine was 11.2%. (8).

The low quality of food in public hospitals and the fact that hospital staff is eating this type of food daily can cause migraines to worsen, so the clinical manifestations of migraine are also affected by nutritional and gastrointestinal factors (9). According to this fact, migraine is a benign headache, but migraine headaches can cause significant disturbances in daily functioning and social interactions. (10)

Research and clinical evidence show that early life stress contributes to long-term changes in the sympathetic nervous system, which are the main pathways for anxiety. (11)

Multiple studies have examined the relationship between migraine headaches and physical fatigue, emotional stress, depression, and other factors in the general population and specific subgroups, including students (12). According to the study Yamanaka et al. (13) showed that behavioral and lifestyle interventions are not proven to reduce headaches. Patients should avoid specific triggers, such as stress and sleep disorders, and make a few diet changes.

The etiology of migraine has been the subject of many hypotheses, but the main cause still remains unknown. Migraine attacks can be prevented by avoiding the factors that lead to their onset or exacerbation.

Since migraine headaches are diagnosed using the International Society of Headache diagnostic criteria, and there is no test to diagnose them (14), hospitals can be a potentially dangerous and stressful environment that can exacerbate migraine headaches and affect social and personal life. This study aimed to examine the prevalence of migraine headaches among the staff of Shahid Beheshti Hospital in Kashan, as well as the relationship between migraine and a variety of factors that it may increase the level of social and individual functioning.

## Method

### Study design

This is a descriptive and cross-sectional study that was carried out among all staff at shahid Beheshti hospital, Kashan of Iran in 2019.

### Subjects

Using a Census method with a sample size of 1125 people was performed in 2014. Participants were staff of Shahid Beheshti hospital, aged between 19 and 62 years.

## Measurements

### Questioners

First, a two-person team surveyed actual hospital staff (1125 people), and then, we use two questinure for this study.

The first questionnaire included demographic questions related to the patient (questions on variables of socio-cognitive population, age, gender, occupation(The hospital employees are divided into five groups (faculty, paramedical, administrative, services, and nurses), and the second job, work shift and sleep hour per day, headache history.

The second questionnaire consisted of two parts, the first part was based on IHS criteria (International Association on Pain (whose reliability and validity are confirmed), which includes 10 questions and identifies the existence or non-existence of migraine.

Those who did not have pain or intervals of more than three months were considered without migraine. If necessary, paraclinical examinations, such as brain scans, were performed to rule out other causes of pain.

Finally, people with migraines were given the second part of the second questionnaire, which includes the clinical features of the disorder (including the average frequency of migraine attacks per month, pain side, photophobia, phonophobia, and visual or sensory aura). Participants were met by the researcher at work and some at home, and the purpose of the study was explained to them and they were informed about the voluntary partnership. The participants completed questionnaires, and if they were unable to do so, interviewers read the questions to them and recorded their responses.

### Ethical consideration

In this study to adhere to ethical considerations, the researcher obtained permission from Kashan Research Ethics Committee(IR.KAUMS.MEDNT.REC.1396.70). Also, the researcher got informed consent from subjects before data gathering and all information of participants was secure.

### Statistical analysis

The questionnaire information was entered into the SPSS software and charts and tables were plotted using Excel software. If needed, Chi-square and Fisher’s test were used. Moreover, the Mann-Whitney test, Kolmogorov-Smirnov test, and Chi-square test were used. Odds of ratio (OR0) were also calculated. Given the 10% prevalence of the migraine in the community (17).

## Results

Among the 1125 staff invited to participate, 58 declined to complete the survey, and 43 submitted incomplete questionnaires. The response rate was 90%. 1022 respondents completed the survey. The age ranged from 18-65years with a median of 29 ±3.2 years.

A study of 1022 people found that only 136 (12.3%) were diagnosed with migraines, 255 people had primary headaches, and a total of approximately 886 people were not diagnosed with migraine based on the HIS criteria. Based on the results, the highest prevalence of migraine was observed in nurses (63 patients, 14.5%) and the lowest in faculty members (6 patients,9.7%). (Table 1)

**Table-1:** The frequency of migraine headaches by occupation

| migraine |  | nurse | services | administrative | paramedical | faculty | total |
| --- | --- | --- | --- | --- | --- | --- | --- |
| Yes | number | 63 | 35 | 21 | 11 | 6 | 136 |
|  | percent | 14.5 | 10.9 | 18.4 | 6.3 | 9.7 | 12.3 |
| No | number | 287 | 287 | 93 | 163 | 56 | 886 |
|  | percent | 85.5 | 89.1 | 81.6 | 93.7 | 90.3 | 87.7 |
| total | number | 350 | 322 | 114 | 174 | 62 | 1022 |
|  | percent | 100 | 100 | 100 | 100 | 100 | 100 |

In terms of gender, women with 93 cases (in women with a mean age of 31 years was higher) (15.5%) had a higher prevalence than men with 43 cases (8.5%). The prevalence of migraine was not significantly related to the second job (920 people did not have a second job, and only 102 people had a second job) (PV = 0.086).In terms of marital status, 94 migraine patients were married and 42 were single with PV = 0.001.

Prevalence of migraine by pain location in 122 patients (89.7%) with unilateral pain and other pain sites are shown in the table below (Table -2)

**Table 2.** Frequency distribution of migraine headache by pain location

| Pain location | frequency | percent |
| --- | --- | --- |
| unilateral | 122 | 89.7 |
| bilateral | 8 | 5.9 |
| Front of head | 5 | 3.7 |
| Back of head | 1 | 0.7 |
| number | 136 | 100 |

The highest frequency of attacks was found to be five per month (32.35%) and the lowest was three attacks with a frequency of one (0.73%). There were 74 cases of anorexia among females (79.6%), compared to 40 cases in men with PV = 0.048.Additionally, nausea was reported to be significant in 77.4% of female (n = 72) and 47% of male (n = 42) with PV = 0.003 (Table-3).

**Table 3.** The most common migraine symptoms

| Common migraine | gender | N(percent) | P-Value |
| --- | --- | --- | --- |
| Anorexia | male | 40(93%) | 0.048 |
|  | Female | 74(79.6%) |  |
| nausea | Male | 42(97.7%) | 0.003 |
|  | female | 72(77.4%) |  |

No significant relationship was found between gender and vomiting PV = 097 and fear of light PV = 0.66, sound PV = 0.13 and odor PV = 1.The frequency of migraine was significantly related to the age of migraine with a mean of 31.07, PV = 0.035, and the average sleep time in people with migraine were 6 hours per day with PV = 0.012.

Out of 136 people with migraine, only 57 had all three clinical signs of migraine (anorexia, nausea, and vomiting), 74 of them worked night shifts and 62 people had morning shifts that had no significant relationship with migraines (PV = 0.5).

## Discussion

Early headaches can affect anyone at any age(15). We included participants who even had a history of one-time headaches because this type of question increased our screening sensitivity in diagnosing migraines, which is why about 25% of participants reported headaches. Because we used the International Headache Association criteria to identify migraine cases in the next step, we did not examine staff with other headaches.

We conducted a population-based study of migraine headaches at the Shahid Beheshti Hospital in Kashan, where migraine prevalence was 12.3%.

### A Comparison of migraine prevalence with other studies and influence on related factors

#### Mean age

Among our study participants, 15.5% were female, and 85% were male, indicating a high prevalence of migraine in females, in similar studies conducted by Dr. Hamzei Moghadam et al. (17), out of 1,029 people (473 women and 556 men) employed in government agencies, 10.4% had migraines according to HIS criteria, of which 11% were female, and 89% were male. The mean age of all people with migraines was 34 years. Showed a significant relationship between age and sex. that was consistent with the results of our study.

in contrast, our study, In the study of Mohdreza Shahraki et al. (18), which examined the prevalence of migraine among 1539 school teachers in Zahedan, no correlation was found between the prevalence of migraine and age, and a similar study by Tayebeh Khazaei et al. (19), they studied the prevalence of migraine among 1107 high school children in Birjand, which, found no significant relationship with age and sex.

In another study, the prevalence of migraine among medical students in Bandar Abbas who are active in the hospital environment was equal to 16.3%, the prevalence was different according to gender, which was higher among male students.(20)

### Sleep hour per day

In another study conducted by Ayatollah et al. (21) to determine the prevalence of tension headaches and migraine headaches among Shiraz university students, 54 (10%) reported migraines that showed a significant relationship with sleep(PV = 0.01). In this study (21), migraine prevalence and sleep are related (PV = 0.012). The number of disability migraine is higher among married people than among singles, according to our research.

In our study, migraine sufferers slept an average of six hours per day. In a separate study. In a similar study, sleep disturbance was also identified as one of the most common headache triggers, which was consistent with our findings, that those who slept an average of six hours a day had more frequent and severe headaches than those who slept more, as well as more headaches in the morning.(22)

### Other factors

Out of 764 patients admitted to Isfahan Neurology Clinic, Chitsaz (23) studied the epidemiological and clinical features of migraine headaches among patients. it was reported only 11% them suffered from migraine headaches. Fifty-five percent of the patients were females and forty-five percent were males. The mean age of migraine patients was 31.3 years and 60.7% of them were married.(similar to our study), Stress (73.8%), sleep disorders (61.9%), and fatigue (59.5%) were the most common migraine headache triggers. The mean number of headache attacks per month was 8.2, and the average duration of each headache attack was 14.2 hours. Most patients had unilateral headaches that were consistent with migraines. In our study, The prevalence of migraines was higher in women (15.5%) and lower in men (8.5%). Married people (94 out of 136) also had the highest migraine incidence and average rate of 5 attacks per month with a duration of 24.3 hours. In the evening, migraine prevalence was highest (54.8%), and during the night was lowest (26.2%).

### B The impact of migraines on the workplace and personal life

According to other studies, migraines are affected by a person’s position or type of work. Due to the frequent changes in their work schedules with sleepless nights, hospital workers are more susceptible to migraine as a result of working several shifts in the hospital(24), in our study hospital staff are more likely to suffer from headaches for the following reasons, 1: stressful environment 2: The long working shift 3: present female staffs more than male staff in the hospital

In Emina Sokolovic’s study, which looked at the prevalence of migraines among Swiss hospital staff, the prevalence of migraines, as in our study, was higher among administrative and medical technicians than among physicians. In our study, first nurses(94 patients) and then service departments(35 patients), and the administrative (21 patients) were mostly people with migraines, which is confirmation of our study, It is possible that migraines are more prevalent in hospital settings since stress is a frequent trigger factor for migraine attacks. (25)

## Conclusion

Migraine disorders deserve more attention, which can be reduced by examining the frequency, and identifying a possible risk factor, frequency of attacks, duration, and severity.

On the other hand, following its impact on social life, it can play a crucial role in improving a person’s life and the need to present an effective strategy in a stressful hospital environment.

## Data Availability

all data are not availiable

